# Global patterns of rebound to normal RSV dynamics following COVID-19 suppression

**DOI:** 10.1101/2024.02.23.24303265

**Authors:** Deus Thindwa, Ke Li, Dominic Cooper-Wootton, Zhe Zheng, Virginia E Pitzer, Daniel M Weinberger

## Abstract

**Introduction:** Annual epidemics of respiratory synctial virus (RSV) had consistent timing and intensity between seasons prior to the SARS-CoV-2 pandemic (COVID-19). However, starting in April 2020, RSV seasonal activity declined due to COVID-19 non-pharmaceutical interventions (NPIs) before re-emerging after relaxation of NPIs. We described the unusual patterns of RSV epidemics that occurred in multiple subsequent waves following COVID-19 in different countries and explored factors associated with these patterns.

**Methods:** Weekly cases of RSV from twenty-eight countries were obtained from the World Health Organisation and combined with data on country-level characteristics and the stringency of the COVID-19 response. Dynamic time warping and regression were used to describe epidemic characteristics, cluster time series patterns, and identify related factors.

**Results:** While the first wave of RSV epidemics following pandemic suppression exhibited unusual patterns, the second and third waves more closely resembled typical RSV patterns in many countries. Post-pandemic RSV patterns differed in their intensity and/or timing, with several broad patterns across the countries. The onset and peak timings of the first and second waves of RSV epidemics following COVID-19 suppression were earlier in the Southern Hemisphere. The second wave of RSV epidemics was also earlier with higher population density, and delayed if the intensity of the first wave was higher. More stringent NPIs were associated with lower RSV growth rate and intensity and a shorter gap between the first and second waves.

**Conclusion:** Patterns of RSV activity have largely returned to normal following successive waves in the post-pandemic era. Onset and peak timings of future epidemics following disruption of normal RSV dynamics need close monitoring to inform the delivery of preventive and control measures.

## Introduction

Respiratory syncytial virus (RSV) causes a high burden of respiratory disease among infants and older adults [1–3] and is an important cause of death in the first year of life [1]. RSV spreads through contact with infectious droplets, and transmission is strongly seasonal, with annual epidemics in most regions typically occurring during the autumn or winter seasons [4]. Understanding temporal variations in healthcare utilization due to RSV is vital to inform planning of hospital capacity, administration of immunoprophylaxis, timing of vaccination, and enrollment for clinical trials of RSV prevention and treatment [5].

Prior to the pandemic caused by SARS-CoV-2 (COVID-19), annual epidemics of RSV largely followed consistent temporal patterns year-to-year in each country [6]. However, during the COVID-19 pandemic, RSV seasonal activity decreased to very low levels starting in March 2020 [7–9]. This was likely due to implementation of non-pharmaceutical interventions (NPIs) such as country-wide lockdowns, border closures, social distancing, face masks, and school closures that were intended to contain the spread of SARS-CoV-2 [7, 10, 11]. After the early phase of the COVID-19 pandemic and the easing of some NPIs [12], RSV activity increased dramatically and heterogeneously across countries. The characteristics of these resurgent epidemics were unexpected and included out-of-season epidemics, a high volume of cases, and multiple within-country outbreaks [13–16]. These unusual patterns highlighted the current and future need to better understand RSV epidemic patterns.

A number of factors were associated with the timing of the initial rebound in RSV activity following COVID-19 suppression [11, 17]. Reopening of schools, increasing population susceptibility or “immunity debt”, and decreasing temperatures were reported to increase the risk of initial RSV rebound [10, 11, 17]. However, as predicted by modeling studies during the initial period of disruption [18], it could take several seasons to return to pre-pandemic seasonal patterns of RSV activity (“return to normal”). The determinants of the return to normal are not fully understood.

In this study, we collated epidemiologic, demographic, climate, geography, and behavioral data from twenty-eight countries globally, from 2017 to 2023. We used these data to measure the correlation of RSV patterns between pre- and post-COVID-19 pandemic period, identify distinct patterns of RSV related to onset, peak timing, growth rate and intensity, and explore their associated factors following RSV disruptions related to COVID-19.

## Methods

### Ethics statement

These analyses used publicly available aggregate time series data at the country level and thus did not contain any data on individual human subjects. These analyses followed the guidance for the Conduct and Reporting of Modeling and Simulation Studies in the Context of Health Technology Assessment [19].

### Data description

We obtained publicly available data on weekly counts of RSV cases by country and hemisphere from January 2017 to July 2023 from the World Health Organisation (WHO) FluNet platform [6, 20], which is a web-based tool for influenza and RSV virological surveillance (https://frontdoor-l4uikgap6gz3m.azurefd.net/FLUMART/VIW_FNT?$format=csv) [6]. We included countries which met the criteria of reporting at least 100 annual RSV cases between 2017 and 2023. In brief, RSV data were transmitted to FluNet from each country’s recognized national influenza center or national public health laboratory with ongoing influenza surveillance and laboratory capacity for RSV testing using molecular methods and with a history of successful WHO external quality assessment for the molecular detection of influenza [6]. Additional data on a country’s climate zone were obtained from the US National Oceanic Atmospheric Administration based on Köppen and Geiger classification [21], whereas a country’s COVID-19 contact stringency index and population density were obtained from the Oxford COVID-19 Government Response Tracker [12]. Population density was calculated as the number of people per square kilometers of land area in 2020 in each country. Stringency index was calculated as a composite measure of the government’s response based on nine response indicators including school closures, workplace closures, and travel bans, rescaled to a value from 0 to 100, (100 = strictest).

### Statistical analyses

#### Dynamic time warping and time series classification

We sought to cluster RSV time series based on the dynamics during the pre- and post-pandemic periods in 28 countries. To do this, we used dynamic time warping (DTW) to evaluate pairwise similarity between time series. We focused on the period from Jan-08-2017 to Dec-31-2022 in the Southern Hemisphere and from Jun-11-2017 to Jun-04-2023 in the Northern Hemisphere [22]. DTW, as implemented in the dtwclust and dtw R packages, aligns a query and reference pair of time series based on common features, and calculates distances between the two time series to form a local cost matrix (lcm) [22–24]. A warping distance, which minimizes the alignment between the two time series, is computed under the constraint of Sakoe-Chiba band window size of the lcm. We selected an optimal window size from a range of values through evaluation of hierarchical clusters as detailed in the supplement [24–26] (Figure 1, Text S1). Hierarchical clustering was performed based on the estimated warping distances [27] and was visualized as a dendrogram. A prototype time series for each cluster summarized the most important characteristics of all series in each cluster using a DTW barycentre averaging function [24]. We conducted sensitivity analysis to assess the alternative number of clusters and DTW window size.

**Figure 1.**
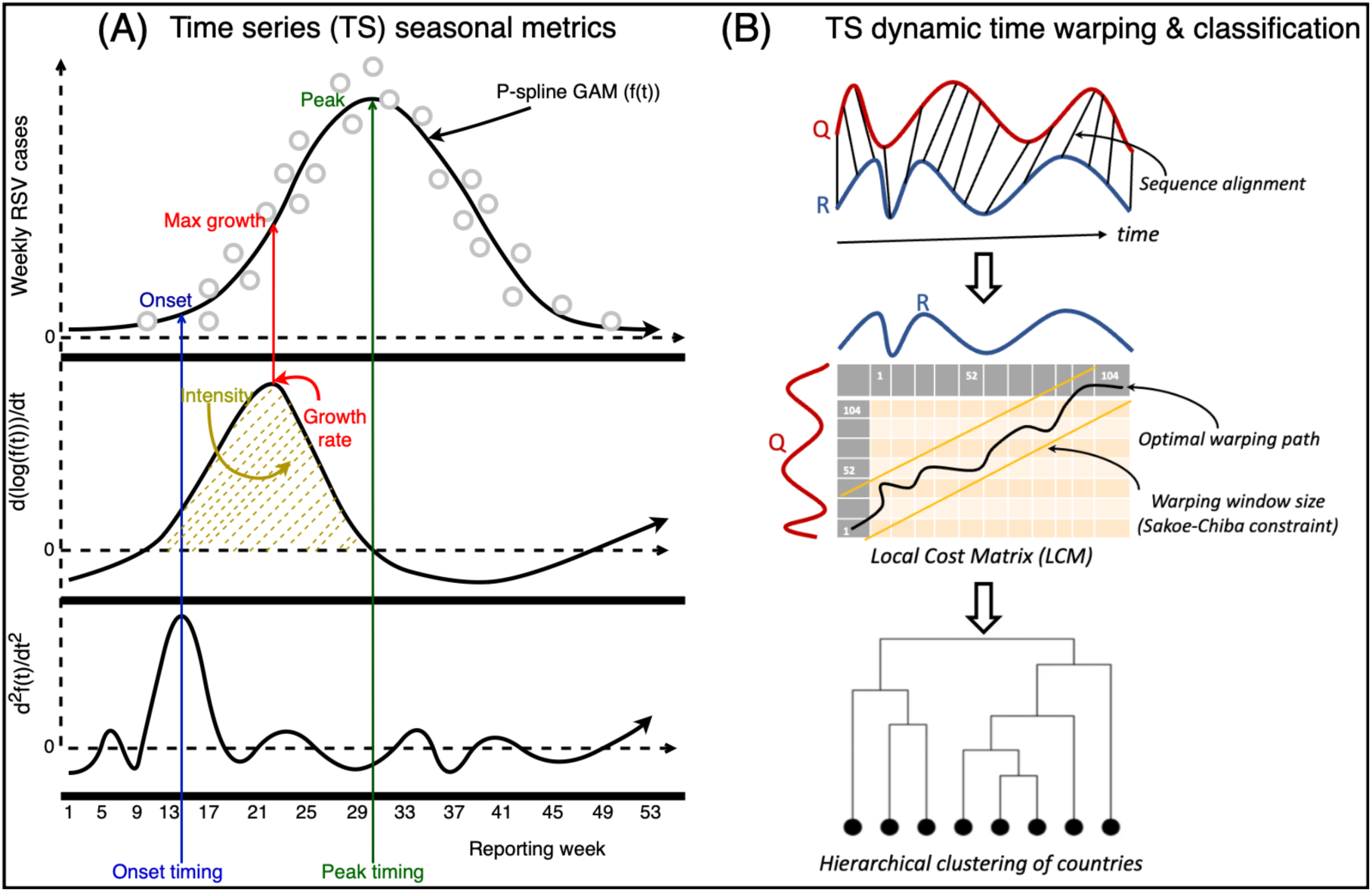
Schematic diagram of respiratory syncytial virus (RSV) seasonal metrics and time series classification. (A) A P-spline (black line curve) is fitted to weekly RSV case counts (circles); peak timing is the week of maximum number of seasonal cases corresponding to the week when the fitted P-spline curve in each season/wave is at a maximum (A-top). The growth rate is the weekly change in new cases corresponding to the maximum value of the derivative of log fitted P-spline curve; and intensity is the fraction of cases before the epidemic peak, corresponding to the integral of positive segment of first derivative of the log fitted P-spline curve (A-middle). Onset timing is the week when the growth rate increases the most, corresponding to week when second derivative of the fitted P-spline curve is at a maximum (A-bottom); (B) Dynamic time warping (DTW) of RSV time series (TS); the time series of any two countries are realigned to measure their dissimilarity (B-top), based on optimal warping distance from a local cost matrix (B-middle), resulting in a hierarchical clustering of countries (B-bottom). Created with Draw.io software [42].

#### Description of post-pandemic epidemic patterns

Since some epidemics in the Southern (e.g., South Africa, Australia) and Northern (e.g., United States, the Netherlands) hemispheres occurred during unusual times of the year following COVID-19 suppression, we report on metrics describing the first, second and third waves of RSV in comparison to a ‘typical season’ from the pre-COVID-19 era. “Epidemic years” are defined as weeks 1-52 of the calendar year for the Southern hemisphere and weeks 24-52 and 1-23 of successive calendar years for the Northern hemisphere. The three waves were defined as separate epidemics in a given country if their onsets occurred at least eight weeks apart.

We computed four metrics to describe the dynamics of RSV activity following suppression of transmission during the initial part of the COVID-19 pandemic (starting April 1, 2020): RSV onset timing, peak timing, growth rate, and intensity. To estimate these metrics, we fitted generalized additive models (GAM) with penalized B-splines (P-splines) to weekly counts of RSV cases over the entire 312-week study period, as defined separately above for the Northern and Southern hemispheres [28]. Details of the GAM P-spline model are presented in the Supplement (Figure 1, Text S2).

Using the fitted P-spline curves, we estimated the following metrics for each pre-COVID-19 season and post-COVID-19 wave [29]:

a. Onset timing: the week when the growth rate increased the most, corresponding to the time point when the second derivative of the P-spline curve was at the maximum value during the period when the growth rate (first derivative) was increasing in each season or wave (Figure S1).
b. Peak timing: the week with the highest number of cases in each season or wave, corresponding to the highest points of the fitted P-spline curve (Figure S2).
c. Growth rate: the rate of change of log-tranformed cases, corresponding to the maximum value of the derivative of the log-transformed fitted P-spline curve in each season or wave (Figure S3).
d. Intensity: the relative magnitude of cases before the epidemic peak in each season or wave, corresponding to the integral of the positive derivative of the log-transformed fitted P-spline curve (Figure S4).

#### Predictors of post-pandemic patterns

For each metric described above, we compared the pre-pandemic RSV seasonal patterns with those observed in the first, second and third RSV epidemic waves in various countries following COVID-19 suppression. We used Pearson’s correlation coefficients to quantify relationships between the covariates and RSV growth rate and intensity and circular correlation coefficients to quantify relationships with RSV onset and peak timing, stratified by hemisphere and climate zone.

We used Cox Proportional Hazard regression to evaluate potential predictors of the time to onset or time to peak of the first epidemic wave after April 2020. We also evaluated predictors of the time between onset or peak timing of the second epidemic wave and the onset of the first wave. We further evaluated predictors of RSV growth rate and intensity in each post-pandemic wave using linear regression. Potential predictors included hemisphere, climate zone, contact stringency index, population density, and out-of-season status and intensity during first RSV wave. All potential predictors were included in univariate analyses, and those with p<0.10 were retained for multivariate analyses. Univariate predictors were included in the multivariate models through a stepwise forward selection procedure if they reduced the Akaike Information Criterion (AIC) score [30, 31].

## Results

### Data Description

There were 28 countries that met the inclusion criteria and were included in the analysis. Of these, 22 (78.6%) and 6 (21.4%) were from the Northern and Southern hemispheres, respectively. Thirteen (46.4%) countries had temperate climates, and 15 (53.6%) were from (sub)tropical zones, with representation from Europe (n=12, 42.9%), South America (n=6, 21.4%), North America (n=3, 10.7%), Western Pacific (n=3, 10.7%), Africa (n=1, 3.6%), Southeast Asia (n=1, 3.6%), and Eastern Mediterranean (2, 7.1%). Population density and median age were higher in the Northern than Southern hemisphere, and in the temperate than (sub)tropical climate zones (Figure S5). Contact stringency index largely declined from 2020 to 2023, with varying rates of decline between countries (Figure S6). While most countries had two RSV waves for the post-pandemic period of our analysis, Australia, Brazil, Colombia, France, Iceland, the Netherlands and South Africa had three waves (Figure S1).

### Epidemic patterns were more normal during the second wave

The first epidemic wave following COVID-19 generally occurred during a time of year when RSV was not typically present (Figure 2, Figure 3). Epidemic onset was more than 8 weeks different from normal seasonal timing in 18 (64.3%) countries. However, in the second and third waves of RSV following COVID-19, onset timing was closer to normal across all countries (circular correlation coefficient of *c*_3_ = 0.99, *c*_2_ = 0.64, *c*_1_ = 0.08 in the third, second, and first waves, respectively) (Figure 3). This was largely seen in both the Northern hemisphere (*c*_3_= 0.99, *c*_2_ = 0.50, *c*_1_ = 0.09) and Southern hemisphere (*c*_3_= 1.00, *c*_2_ = 0.25, *c*_1_ = 0.46). Similar patterns were seen when evaluating peak timing across all countries (*c*_3_= 0.43, *c*_2_ = 0.16, *c*_1_ = 0.14), in the Northern hemisphere (*c*_3_= 0.45, *c*_2_ = 0.13, *c*_1_ = 0.19), and in the Southern hemisphere (*c*_3_ = 0.94, *c*_2_ = 0.55, *c*_1_ = 0.63). Similar movement towards normal dynamics was also seen when comparing growth rates and intensity in both the Northern and Southern hemispheres (Figure S7) and across climate zones (Figure S8). Among countries with three waves only, correlation estimates of RSV onset, growth rates and intensity were also closer to pre-COVID-19 mean estimates for the third compared to the second wave (Table S1).

**Figure 2.**
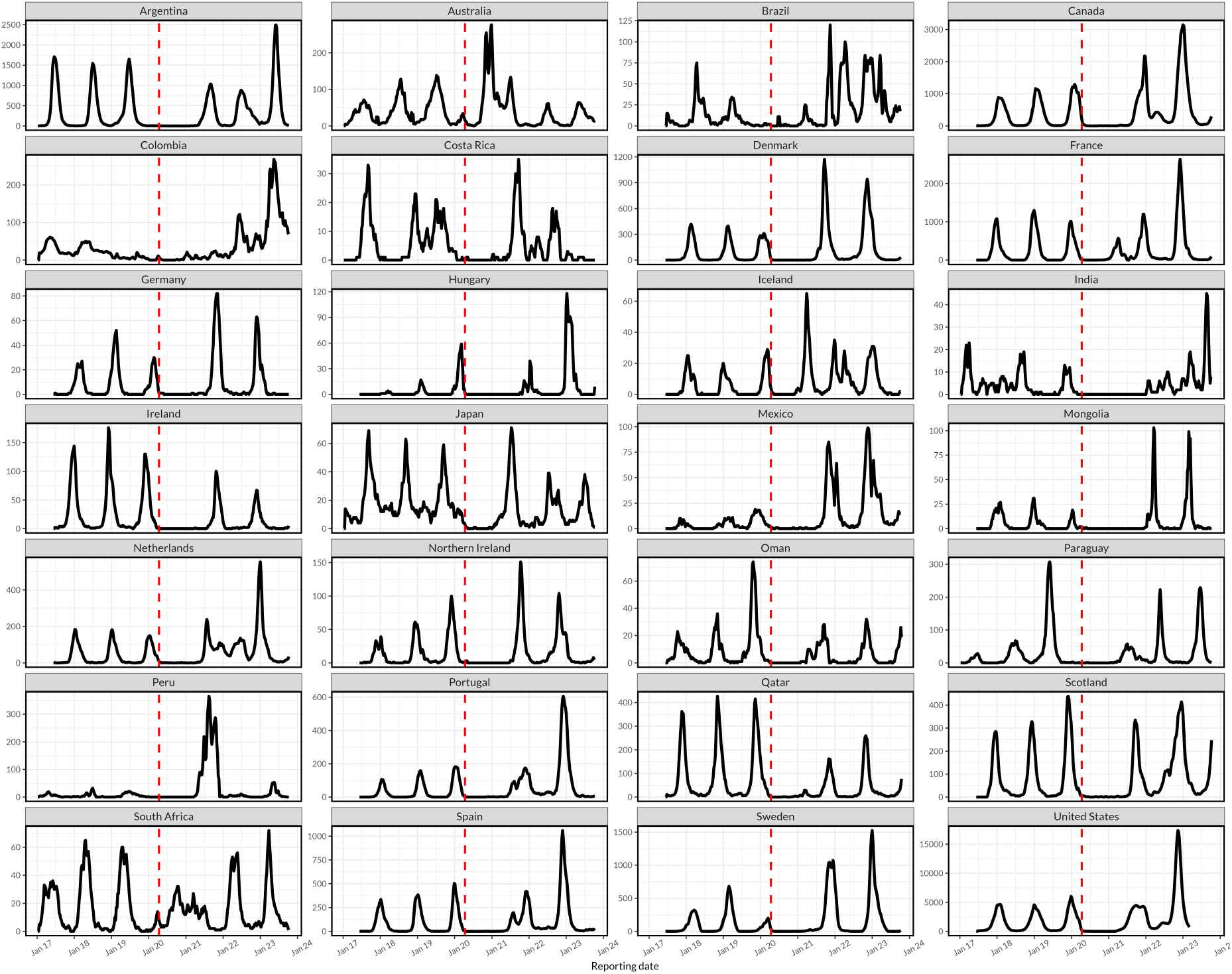
Time series of respiratory syncytial virus (RSV) epidemics across 28 countries. RSV time series from 2017 to 2023 for countries in the Northern and Southern hemispheres show substantial declines in the number of reported cases following the onset of the SARS-CoV-2 pandemic (COVID-19) in 2020 when non-pharmaceutical intenrventions were implemented in most countries (the onset of COVID-19 is shown by a red vertical line). Heterogeneous RSV patterns are observed thereafter, including unexpected delayed onset, elevated intensity, or multiple annual outbreaks within countries when compared to the pre-COVID-19 period.

**Figure 3.**
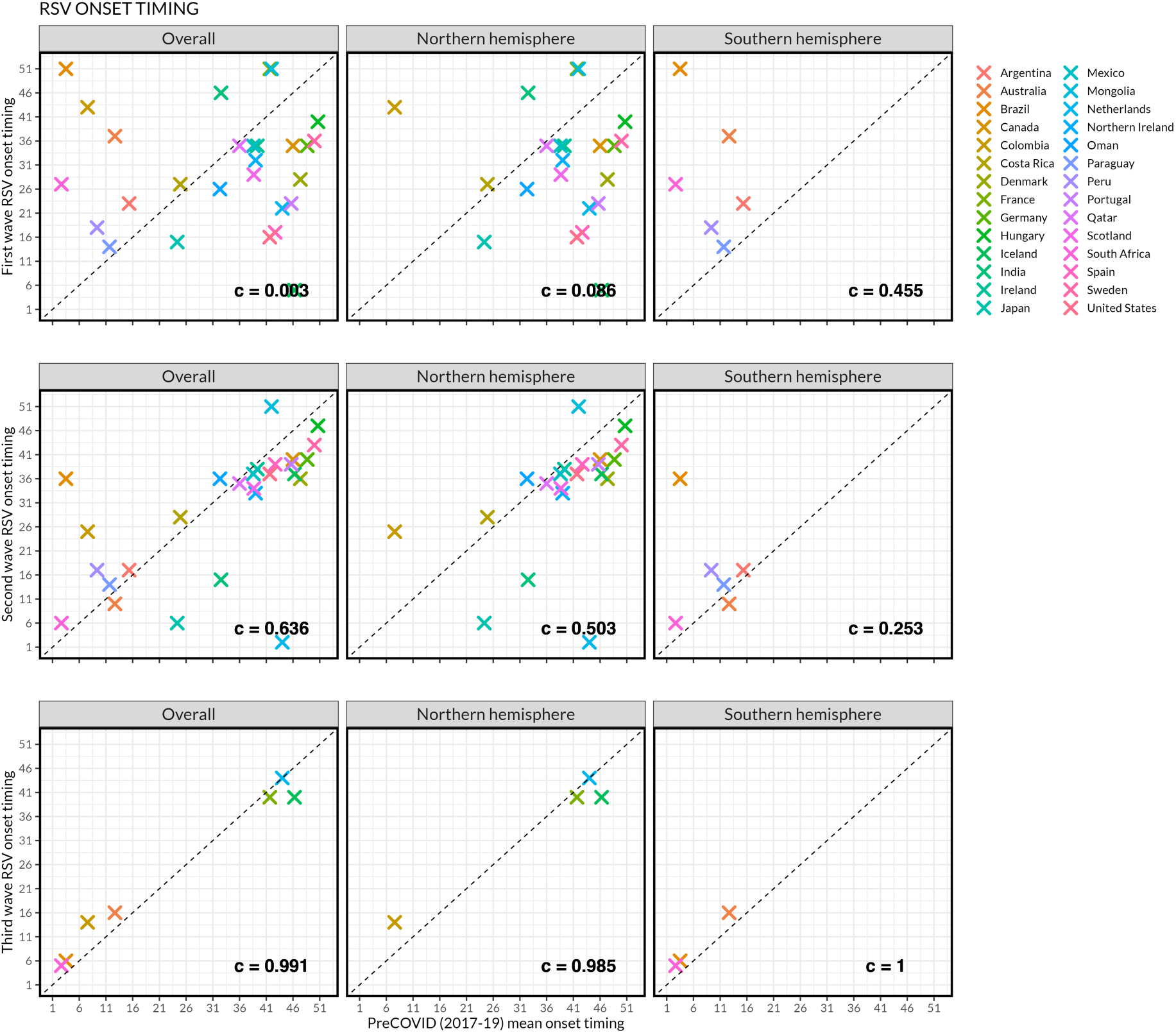
Respiratory syncytial virus (RSV) epidemic onset timing during the first, second and third waves of RSV compared to mean pre-COVID-19 timing in all countries, and by Northern and Southern hemispheres. The *c* metric refers to the circular correlation coefficient. Overall, the plot shows that while the first wave of RSV post-COVID-19 suppression exhibited unusual patterns, the second wave and third wave more closely resembled typical RSV patterns in many countries.

We observed patterns in the relationship between the onset timing and intensity of RSV epidemic waves (Figure 4). Generally, countries with three waves and out-of-season RSV onset (e.g. the Netherlands and Colombia in the Northern hemisphere; South Africa and Brazil in the Southern hemisphere) had large first epidemic waves followed by relatively low-intensity epidemics during the second wave. However, their second wave epidemics poorly matched their typical onset timing (Figure 4). On the other hand, the general pattern of countries with two waves showed that, during the second wave, their onset timing had aligned with a typical RSV onset.

**Figure 4.**
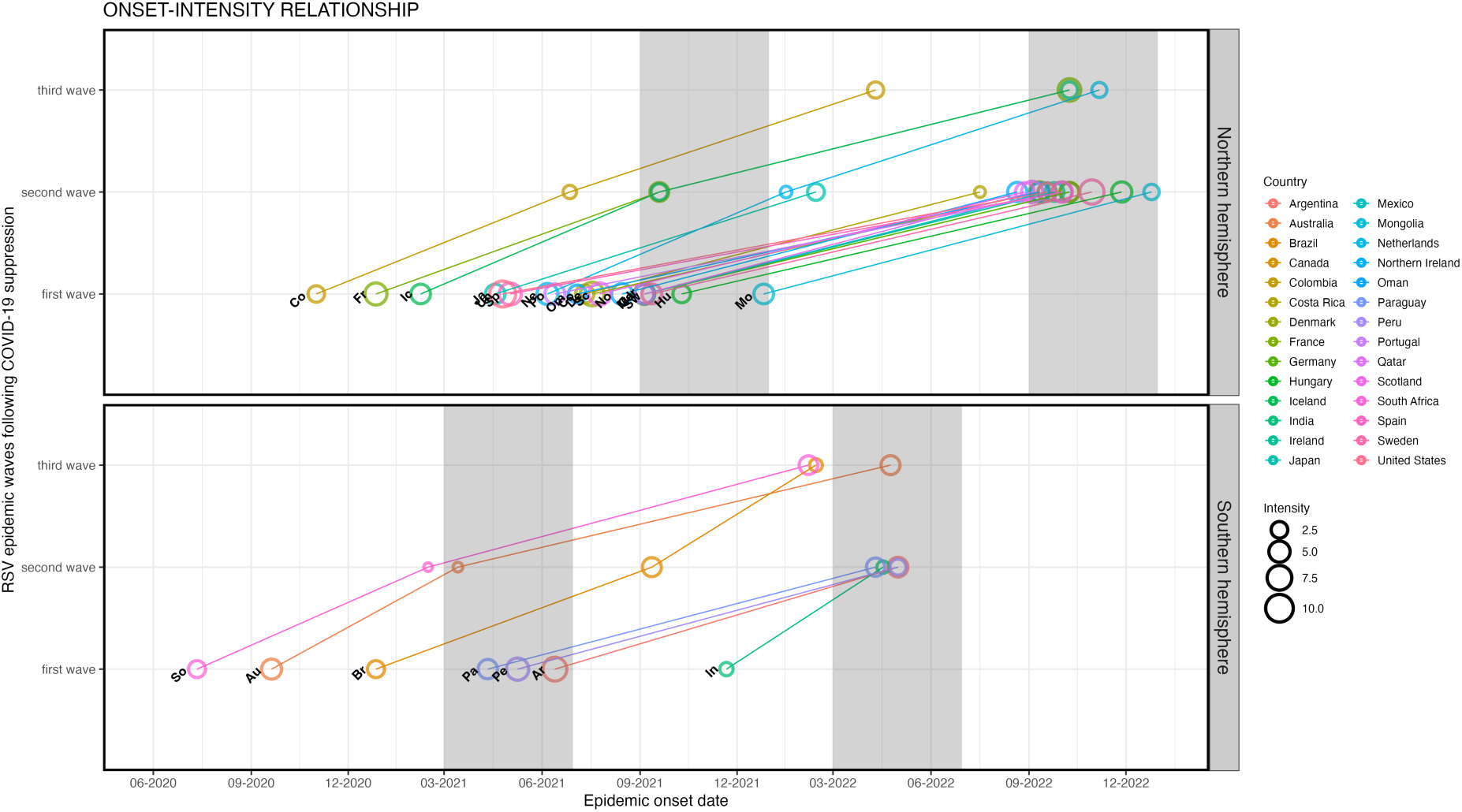
Relationship between respiratory syncytial virus (RSV) epidemic onset and intensity following COVID-19 suppression in April 2020. Epidemic onset and intensity across three RSV waves stratified by hemisphere. Each circle represents a country with RSV onset date on the X-axis and RSV wave on the Y-axis; the size of the circle is proportional to intensity of RSV activity during that wave. The vertical shaded region in each hemisphere represents the onset of a typical RSV epidemic in the pre-COVID-19 period across different countries (e.g. March-June in the Southern hemisphere and September-December in the Northern hemisphere).

### Distinct epidemic clusters during the first and second RSV waves

We identified four clusters of time series, which exhibited four general patterns of RSV activity following COVID-19 suppression (Figure 5). The clusters provide a rough method for grouping countries, and not all countries within a cluster necessarily followed the average patterns; (a) Iceland, South Africa, Qatar, Argentina, Ireland, Oman, and Scotland formed cluster 1, largely characterised by similar delayed and low-intensity patterns of the first and second waves relative to the pre-pandemic era; (b) Australia, Costa Rica, and Japan formed cluster 2, with a high-intensity first wave followed by low-intensity second wave; (c) Brazil, Mexico, Northern Ireland, Mongolia, Germany, Denmark, Sweden, Netherlands, France, Portugal, Canada, Spain, and United States formed cluster 3, with earlier and high-intensity patterns for the first and second waves relative to the pre-pandemic era; and (d) Paraguay, Hungary, and Peru formed cluster 4, with a low-intensity first wave followed by a high-intensity second wave (Figure 5). In a sensitivity analysis, using three clusters (DTW window of size 65) resulted in reclustering of cluster 1 and cluster 3 together, whereas using two clusters (DTW window of size 66) resulted in reclustering of clusters 1, 2, and 3 together (Figure S9, Figure S10).

**Figure 5.**
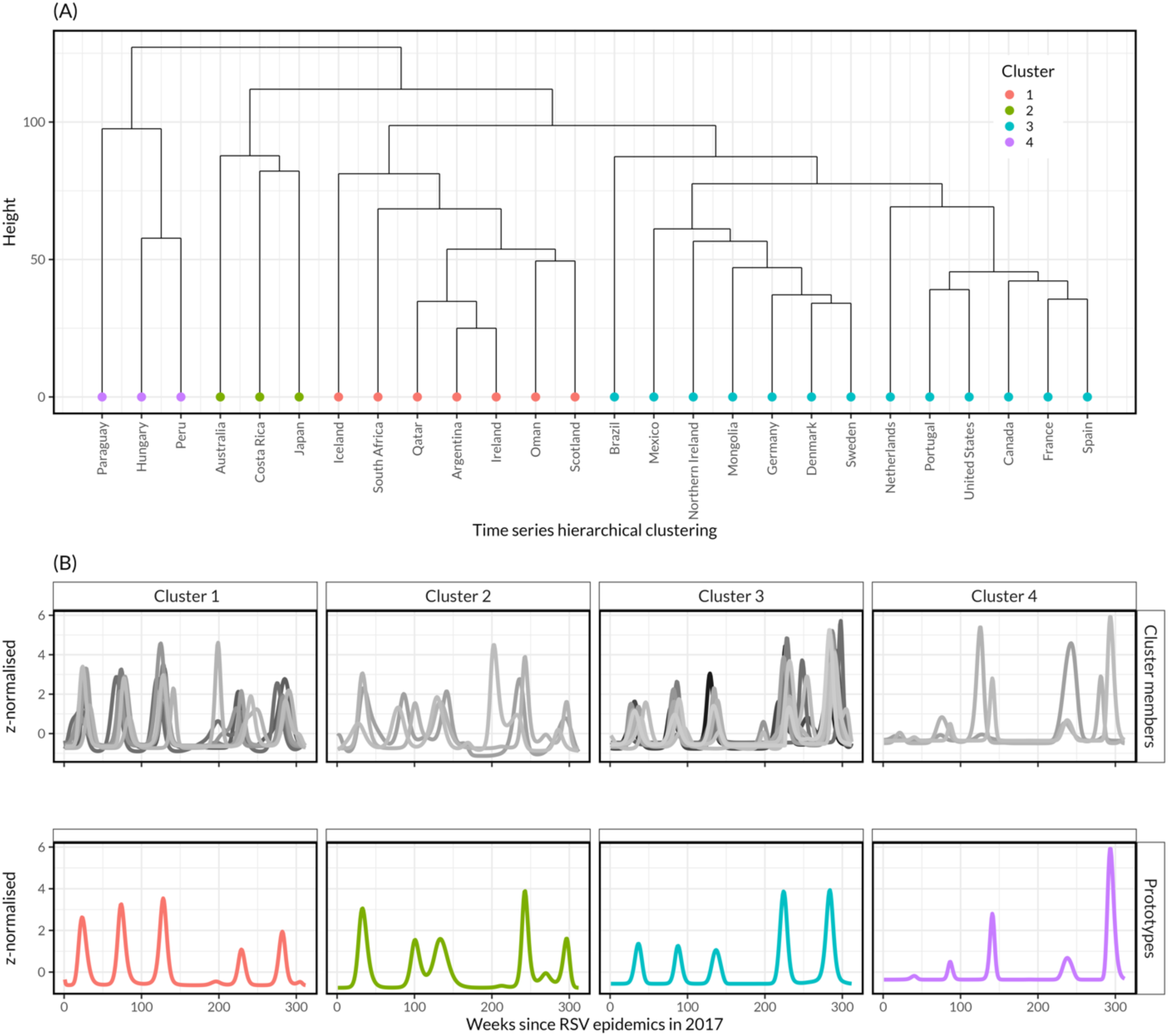
Respiratory syncytial virus (RSV) time series classification of 26 countries using dynamic time-warping (DTW) and hierarchical clustering. (A) Hierarchical clustering of countries based on optimal DTW window size of 74 and 4 clusters, with cluster 1 having 7 countries, cluster 2 having 3 countries, cluster 3 having 13 countries and cluster 4 having 3 countries. (B) Cluster members and their respective prototypes (or centroids from averaging cluster members) based on DTW barycentre averaging. India and Colombia were excluded due to their unique time series.

### Predictors of epidemic characteristics during the first RSV wave

In univariate analysis, during the first wave following COVID-19 suppression, RSV onset and peak timing were earlier in the Southern than Northern hemisphere (hazard ratio (HR): 16.44, 95% confidence interval (CI): 5.06-53.46) and (HR: 20.41, 95% CI: 5.63-73.97), respectively, whereas RSV intensity was lower in tropical/subtropical than temperate areas (0.33, 95% CI: 0.11-1.02) (Table 1).

**Table 1.**
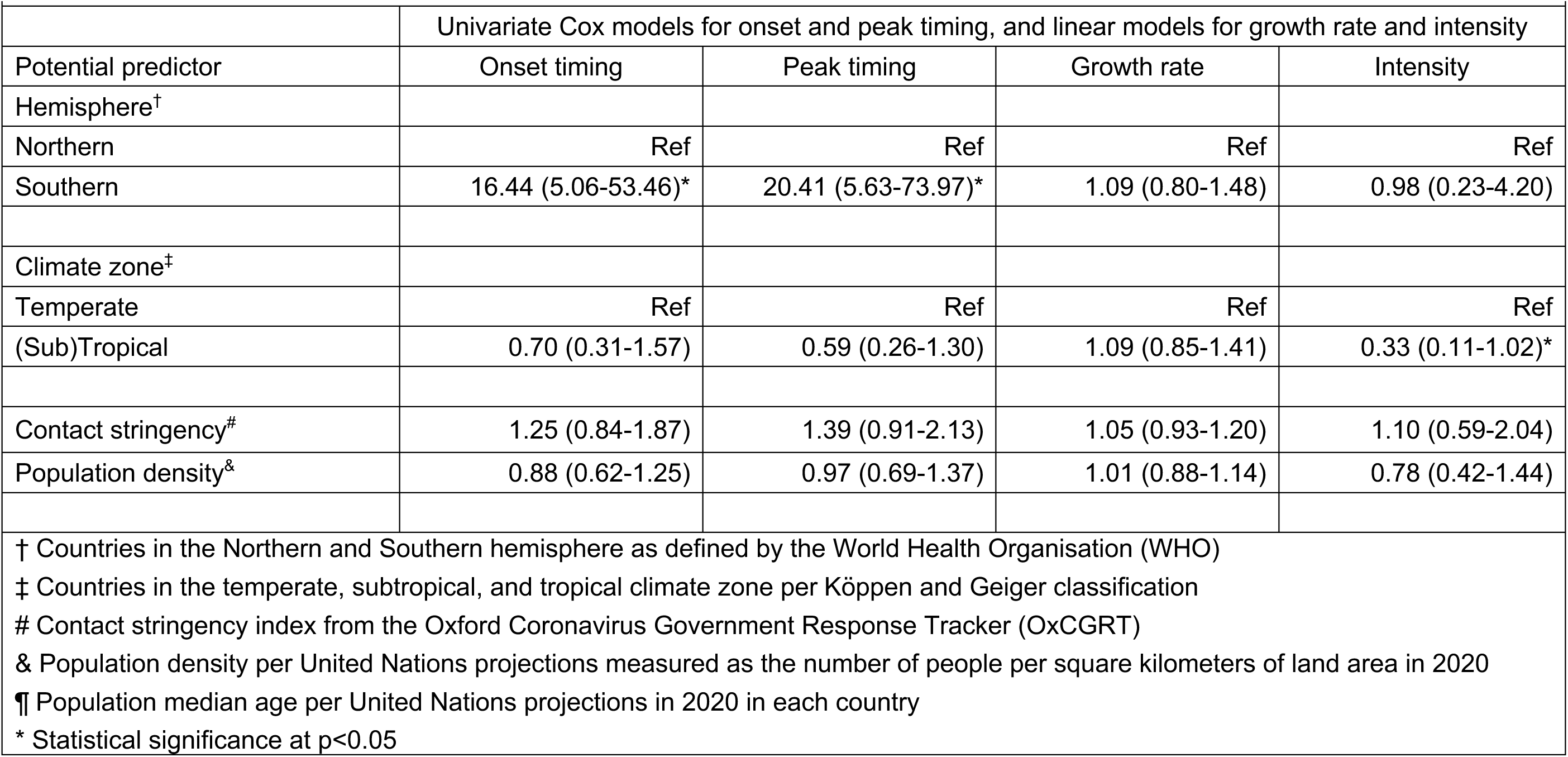
Predictors of respiratory syncytial virus (RSV) onset timing, peak timing, growth rate and intensity during RSV first wave after the early phase of COVID-19 suppression in 2020 using data from 28 countries globally.

### Predictors of epidemic characteristics during the second RSV wave

In multivariate analysis, the gap between the onset of the first and second waves was shorter in the Southern than Northern hemisphere (HR: 13.98, 95% CI: 2.61-74.90) and was earlier in countries with higher contact stringency (HR: 6.47, 95% CI: 2.52-16.62) and higher population density (HR: 3.01, 95% CI: 1.65-5.49). The gap between the onset of the first and second waves was longer with higher first wave intensity (HR: 0.30, 95% CI: 0.17-0.53). Peak timing of the second wave was also earlier in the Southern than Northern hemisphere (HR: 9.36, 95% CI: 2.08-42.20), in the tropical/subtropical than temperate (HR: 3.17, 95% CI: 1.12-8.94) and with higher contact stringency (HR: 3.63, 95% CI: 1.90-6.96) (Table 2).

**Table 2.** Predictors of respiratory syncytial virus (RSV) onset timing, peak timing, growth rate and intensity during RSV second wave following RSV first wave post early phase of COVID-19 suppression in 2020 using data from 28 countries globally.

|  | Univariate Cox models for onset and peak timing, and linear models for growth rate and intensity |  |  |  | Multivariate Cox model for onset and peak timing |  |
| --- | --- | --- | --- | --- | --- | --- |
| Potential predictor | Onset timing | Peak timing | Growth rate | Intensity | Onset timing | Peak timing |
| Hemisphere<sup>†</sup> |  |  |  |  |  |  |
| Northern | Ref | Ref | Ref | Ref | Ref | Ref |
| Southern | 11.13 (3.39-36.58)* | 29.42 (7.28-118.86)* | 0.92 (0.71-1.19) | 0.39 (0.09-1.73) | 13.98 (2.61-74.90)* | 9.36 (2.08-42.20)* |
| Climate zone<sup>‡</sup> |  |  |  |  |  |  |
| Temperate | Ref | Ref | Ref | Ref |  | Ref |
| (Sub)Tropical | 1.00 (0.47-2.15) | 2.46 (1.16-5.20)* | 0.91 (0.73-1.12) | 0.36 (0.11-1.22) | — | 3.17 (1.12-8.94)* |
| Contact stringency<sup>#</sup> | 3.97 (1.96-8.04)* | 3.14 (1.83-5.38)* | 0.88 (0.80-0.97)* | 0.36 (0.22-0.59)* | 6.47 (2.52-16.62)* | 3.63 (1.90-6.96)* |
| Population density<sup>&</sup> | 1.88 (1.12-3.17)* | 0.86 (0.55-1.34) | 0.94 (0.84-1.04) | 0.78 (0.41-1.49) | 3.01 (1.65-5.49)* | — |
| 1<sup>st</sup> wave Intensity | 0.45 (0.28-0.73)* | 0.71 (0.46-1.11) | 0.97 (0.87-1.09) | 1.64 (0.88-3.03) | 0.30 (0.17-0.53)* | — |
| 1<sup>st</sup> wave out-of-season onset<sup>§</sup> | 0.45 (0.19-1.08)** | 0.90 (0.40-2.00) | 1.03 (0.83-1.31) | 0.73 (0.19-2.83) | 0.55 (0.19-1.54) | — |

† Countries in the Northern and Southern hemisphere as defined by the World Health Organisation (WHO)
‡ Countries in the temperate, subtropical, and tropical climate zone per Köppen and Geiger classification

## Discussion

We used time series and regression analyses to understand the global RSV patterns following COVID-19 suppression. While the first post-COVID-19 wave of RSV exhibited unusual patterns, the second and third waves more closely resembled typical RSV patterns in many countries. Post-pandemic RSV patterns were broadly distinct in their intensity and timing of the onset and peak. The onset and peak timings of RSV activity following COVID-19 suppression were earlier in the Southern than Northern hemisphere for both the first and second RSV waves, whereas the intensity of first wave was lower in the tropical/subtropical than temperate countries. The gap between the onset of the first and second waves of RSV was also shorter in the countries with higher population density, and was longer if the intensity of the first wave was higher. While higher contact stringency limited growth rate and intensity, it was associated with a shorter gap between the onset or peak of the first and second waves. Moreover, the peak timing was also earlier in the tropical/subtropical than temperate countries. Overall, our findings underscore different factors that may influence the timing of post-COVID-19 suppression RSV epidemics, which could be useful for anticipating future RSV patterns to inform planning of RSV prevention.

The comparison of epidemic characteristics between the first, second, and third waves of RSV and the pre-pandemic data suggest that RSV patterns have generally returned to typical seasonality. While prior studies have described the initial rebound of RSV following the COVID-19 pandemic [17], it remained uncertain how long it could take to return to normal pre-pandemic seasonal patterns of RSV activity. Our analyses revealed distinct patterns of RSV epidemic characteristics related to timing and/or intensity. There is ongoing interest in the dynamics of RSV during the post-COVID-19 era in different parts of the world [32]; the analyses presented here are updated regularly on an interactive dashboard [33].

Estimated earlier onset and peak timings of post-pandemic RSV waves in the Southern versus Northern hemisphere may likely reflect a higher susceptibility build-up in the Southern hemisphere due to longer time since the last RSV epidemic during pre-COVID-19 period. The association between higher population density and earlier onset of RSV during second wave suggests that countries with more frequent human contacts, due to higher population density, were more likely to experience earlier epidemic starts, consistent with what would be expected based on social mixing patterns and the spread of respiratory pathogens [4, 35, 36].

Interestingly, contact stringency was not associated with the growth rate or intensity during the first wave of RSV following COVID-19 suppression; however, the contact stringency index remained substantially higher (>50% in most countries) and was less variable among countries during this period (Figure S5, Figure S6). However, the substantial reduction and more variability in contact stringency in some countries led to increased RSV growth rate and intensity during the second wave, as shown by our model.

Our model estimated an association between delay of the second wave of RSV and intensity of the first wave. This could be a result of depletion of susceptible individuals and/or generation of sufficient population protective immunity during the first wave [15, 37]. On the other hand, tropical/subtropical countries showed lower intensity during the first wave of RSV, which may have resulted in earlier peaks for the second wave due to availability of enough susceptibles.

Our study had some limitations. Data on the number or percentage of individuals tested for RSV were not available; thus, we could not assess testing or reporting biases in different countries, which may explain some unexpected dynamics in the time series data, e.g. Peru, Mexico, Colombia. To minimise case reporting biases and changes in testing patterns over time, we examined the rate of change of the log-transformed smoothed case counts to represent the relative speed of case accumulation in order to estimate the onset, peak timing, growth rate and intensity for individual epidemic waves [41]. We combined subtropical and tropical climate zone countries due to the small number of available countries in each category. Likewise, we could not perform analyses stratified by WHO regions due to insufficient data from the African and Asian regions. However, available data from other regions such as Europe and Americas were relatively large enough and of good quality. Testing data obtained from WHO platforms may change with time following data cleaning, and our results should be interpreted in the context of the data version at the time of analysis, which are also available in the Github at https://github.com/deusthindwa/rsv.rebound.normal.seasonality.global.

In conclusion, patterns of RSV activity have nearly returned to normal following COVID-19-related suppression. Timing of onset and peak of future RSV patterns need close monitoring to inform the delivery of preventive and control healthcare services.

## Acknowledgements

Research reported in manuscript was fully supported by the National Institutes of Health (NIH) under award number R01AI137093. The content is solely the responsibility of the authors and does not necessarily represent the official views of the National Institutes of Health.

## Author contributions

Conceptualization; DMW, VEP

Data curation; DT

Formal analysis; DT, DMW

Funding acquisition; DMW, VEP

Investigation; DT, DMW

Methodology; DT, DMW, VEP

Project administration; DMW, VEP

Resources; DMW, VEP

Software; DT

Supervision; DMW

Validation; DT, DMW

Visualization; DT, DMW

Writing - original draft; DT

Writing - review & editing; DT, KL, DCW, ZZ, VEP, DMW

All authors read and approved the final manuscript.

## Data availability

The data used in this analysis are publicly available in the GitHub repository (https://github.com/deusthindwa/rsv.rebound.normal.seasonality.global)

