## Supplementary text_tables_figures for "Global patterns of rebound to normal RSV dynamics following COVID-19 suppression"

Supplementary to Thindwa et. al.

**Correspondence**
*
*

### Supplementary Text 1: RSV dynamic time warping and classification

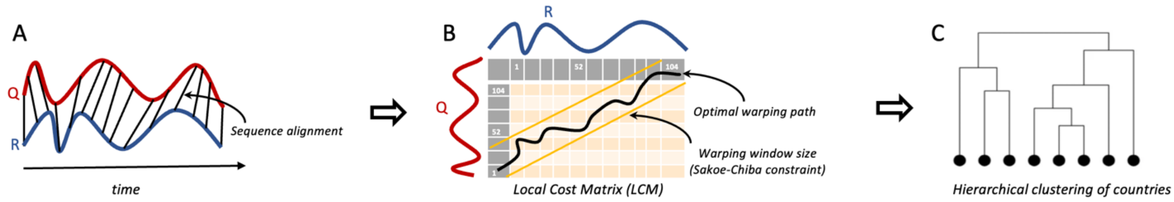


Schematic diagram of dynamic time warping (A), local cost matrix (B) and hierarchical clustering (C) as reference in the text below and drawn using Draw.io software [1]

**Dynamic Time Warping**

We adopted a shape-based time series clustering approach using dynamic time warping (DTW) to quantify dissimilarity between any two time-series (the query-Q and reference-R) from 26 countries, globally [2]. The DTW algorithm computed the optimum warping path (minimum distance) between two time-series under certain constraints including monotonicity, continuity, warping window and boundary. The algorithm initially aligned the two time-series sequences based on common features before computing distances as shown in plot (A). The *dtwclust* and *dtw* R packages facilitated implementation of the algorithm and optimisation [3, 4].

**Local Cost Matrix (LCM)**

After computing distances, a local cost matrix (*lcm*) was created with $n \times m$ dimensions, corresponding to the length of the time series, for every pair of time-series compared. Given the $Q$ and $R$ input time-series, for each element $(i, j)$ of the *lcm* shown as each cell in plot (B), the distance $l_p$ norm (Euclidean) between $Q_i$ and $R_j$ was computed as

$$lcm(i, j) = \left(\sum |Q_i - R_j|^p\right)^{1/p}$$

Hence, the DTW algorithm identified the path that minimizes the alignment between $Q$ and $R$ by iteratively stepping through the *LCM*, starting at $lcm(1,1)$ and finishing at $lcm(n, m)$, and aggregating the cost. At each step, the algorithm found the direction in which the cost increases the least under the above constraints. We defined $\varphi = \{(1,1), \ldots, (n,m)\}$ as a set containing all the points that fell on the optimum path, with the final distance computed as below and facilitated by *proxy* R package [5], where $m_\varphi$ is a per-step weighting coefficient and $M_\varphi$ is the corresponding normalization constant:

$$DTW_p(Q,R) = \left(\sum \frac{m_\varphi lcm(k)^p}{M_\varphi}\right)^{1/p}, \forall k \in \varphi$$

**DTW window size**

To limit the area of the LCM that can be reached by the DTW algorithm to marginally speed up the DTW calculation, we implemented the Sakoe-Chiba window as a global constraint [6], with which the allowed region was created along the diagonal of the LCM as shown in plot (B). For a window size $w$, the valid region of the LCM, the slanted band window, constituted all valid points in the range $[(i, j-w), (i, j+w)]$ for all $(i,j)$ along the LCM diagonal, and at each step, $2w+1$ elements fell within the window. To select an optimal window size and number of clusters for hierarchical clustering of countries as shown in plot (C), we evaluated clustering using Modified Davies-Bouldin (DB) internal cluster validity index (CVI), iterating across different values of window size from 1 to 100 and of cluster size from 2 to 4 [4, 7, 8]. For each window size and cluster size, the DB CVI calculated distances from computed cluster centroids (centroid choice is described below).

**Time series prototype or centroid**

We computed an average series or prototype or centroid to define a time-series that effectively summarizes the most important characteristics of all series in a given cluster. Our choice of prototyping function was the DTW barycentre averaging corresponding to DTW distance measure. The DTW barycentre averaging approach randomly selected one of the series in the data as a centroid, such that on each iteration, the DTW alignment between each series in the cluster and centroid was computed. Warping was performed in DTW, and several time-points from a given time-series mapped to a single time-point in the centroid series, so for each time-point in the centroid, all the corresponding values from all series in a cluster were grouped together according to DTW alignments, and the mean was computed for each centroid point using the values contained in each group. This was iteratively repeated until convergence was assumed (Figure S9).

Hierarchical clustering of countries created a hierarchy of groups in which, as the level in the hierarchy increased, clusters were created by merging clusters from the next lower level, such that an ordered sequence of groupings was obtained [9]. The created hierarchy was visualized as a binary tree using dendrogram where the height of each node was proportional to the value of the inter-group dissimilarity between its two daughter nodes (Figure 4, Figure S10).

### Supplementary Text 2: RSV seasonal metrics calculation

**Generalised additive modelling with P-spline**

Given the time series data from 28 countries, we fitted generalized additive models with penalised B-spline (P-spline) [10]. Weekly RSV cases were assumed to follow a Poisson distribution with mean ($\mu$) as follows.

$y_i \sim Poisson(\mu_i)$,

where $\mu$ is equal to the expectation of $y_i$:

$\mu = \mathbb{E}(y_i)$,

and

$\text{Log}(\mu) = \beta_0 + f(t).$

$\text{Log}\ (\mu)$ is equal to the intercept ($\beta_0$) and smooth function of weekly cases ($f(t)$) using penalized B-splines (P-splines), with a log link function for the Poisson distribution family.

**Epidemic analysis**

We defined four metrics to summarize RSV epidemics before and after COVID-19 suppression period in each of the 28 countries (Figure 1).

1. Onset timing (O) was defined as a week of epidemic start corresponding to the timing of the maximum of the second derivative in the segment of increasing first derivative for the fitted P-spline with respect to time (week); this is mathematically represented as [11]:

$$O = t{:}Max\left\{\frac{d}{dt}\left[\frac{d\big(f(t)\big)}{dt} > 0\right]\right\}$$

2. Peak timing (P) was defined as the week of maximum wave cases, which corresponds to the timing of the maximum value of the fitted P-spline curve in each epidemic wave [12], represented as:

$$P = t{:}Max\{f(t)\}$$

3. Growth rate (G) was defined as the number of new cases per week corresponding to the maximum value of the derivative of the log-transformed fitted P-spline curve with respect to time (week) [13]; this is mathematically represented as:

$$G = Max\left\{\frac{d}{dt}\left[log\big(f(t)\big) > 0\right]\right\}$$

4. Intensity (I) was defined as the relative magnitude of cases before the epidemic peak, corresponding to the integral of the positive derivative of the log fitted P-spline curve with respect to time (week); this is mathematically represented as:

$$I = \int\left\{\frac{d}{dt}\left[log\big(f(t)\big) > 0\right] > 0\right\}dt$$

**Correlation coefficients**

We performed correlation tests between two distinct phases of time series in 28 countries globally to establish degree of return to normal RSV patterns post COVID-19 suppression. We compared pre COVID-19 mean onset and peak timings, growth rate and intensity first wave, second wave and third wave of RSV epidemic following COVID-19 suppression. Onset and peak timings for pre- and post-COVID-19 phases were quantified using circular correlation coefficient ($c$), as implemented in circular R package [14], and visualised using X-Y plots. Assume a sample of $n$ pairs of time series points or angles {$(a_{11}, a_{21})$, $(a_{12}, a_{22})$, …, $(a_{1n}, a_{2n})$} corresponding to the two phases of time series, then the circular correlation is given by:

$$c = \frac{\sum_{k=1}^{n}\sin(a_{1k} - T_{11})\sin\,(a_{2k} - T_{21})}{\sqrt{\sum_{k=1}^{n} sin^2(a_{1k} - T_{11})\sum_{k=1}^{n} sin^2(a_{2k} - T_{21})}}$$

where $T_{11}$ is mean direction of the first circular variable, and $T_{21}$ is the mean direction of the second circular variable.

Growth rate and intensity for pre- and post-COVID-19 phases were quantified using Pearson's correlation coefficient ($r$), as implemented in the stats R package, and visualised using X-Y plots. Assume a sample of $n$ pairs of time series points $\{(x_1, y_1), (x_2, y_2), \ldots, (y_n, y_n)\}$ corresponding to the two phases of time series, then the Pearson's correlation is given by:

$$r = \frac{\sum_{i=1}^{n} (x_i - \bar{x})(y_i - \bar{y})}{\sqrt{\sum_{i=1}^{n} (x_i - \bar{x})^2 \sum_{i=1}^{n} (y_i - \bar{y})^2}}$$

where $\bar{x} = 1/n \sum_{i=1}^{n} x_i$ is the sample mean, and analogously for $\bar{y}$.

**Regression models of RSV onset, peak timing, growth rate, and intensity**

Cox Proportional Hazards regression models were separately fitted to 'time to RSV onset' and 'time to RSV peak' to identify factors associated with the timing of RSV waves, whereas linear models were fitted to 'RSV growth rate' and 'RSV intensity'. Mathematical descriptions of the fully saturated models are given below:

$$Expected\ harzard(time\ to\ onset\ or\ time\ to\ peak\ of\ RSV\ epidemic)$$
$$= (baseline\ hazard) \exp\big(b_1\ \text{Covid19 contact stringency index}_{\text{30 days moving average}}$$
$$+ b_2\ population\ \text{density} + b_3 \text{climate zone} + b_4 \text{hemisphere}$$
$$+ b_5 \text{outOfSeason during RSV 1st wave} + b_6 \text{intensity during RSV 1st wave}\big)$$

where the baseline hazard represents the hazard when all predictors are equal to 0, and $b_1, b_2, \ldots, b_n$ are predictor coefficients. Predictors are included or excluded in the multivariate model based on stepwise selection [15]. The hazard ratio (HR) is the ratio of expected hazards between the two comparison groups.

$$\begin{aligned} \textit{Expected value}(\textit{growth rate or intensity}) \\ &= (\textit{intercept}) + b_1 \text{ Covid19 contact stringency index}_{\text{30 days moving average}} \\ &+ b_2\, \textit{population} \text{ density} + b_3 \text{climate zone} + b_4 \text{hemisphere} \\ &+ b_5 \text{outOfSeason during RSV 1st wave} + b_6 \text{intensity during RSV 1st wave} \end{aligned}$$

where the intercept represents the expected growth rate or intensity when all predictors are equal to 0, and $b_1, b_2, \dots, b_n$ are predictor coefficients. Predictors are included or excluded in the multivariate model based on stepwise selection [15]. The effect size is the ratio of expected values between the two comparison groups.

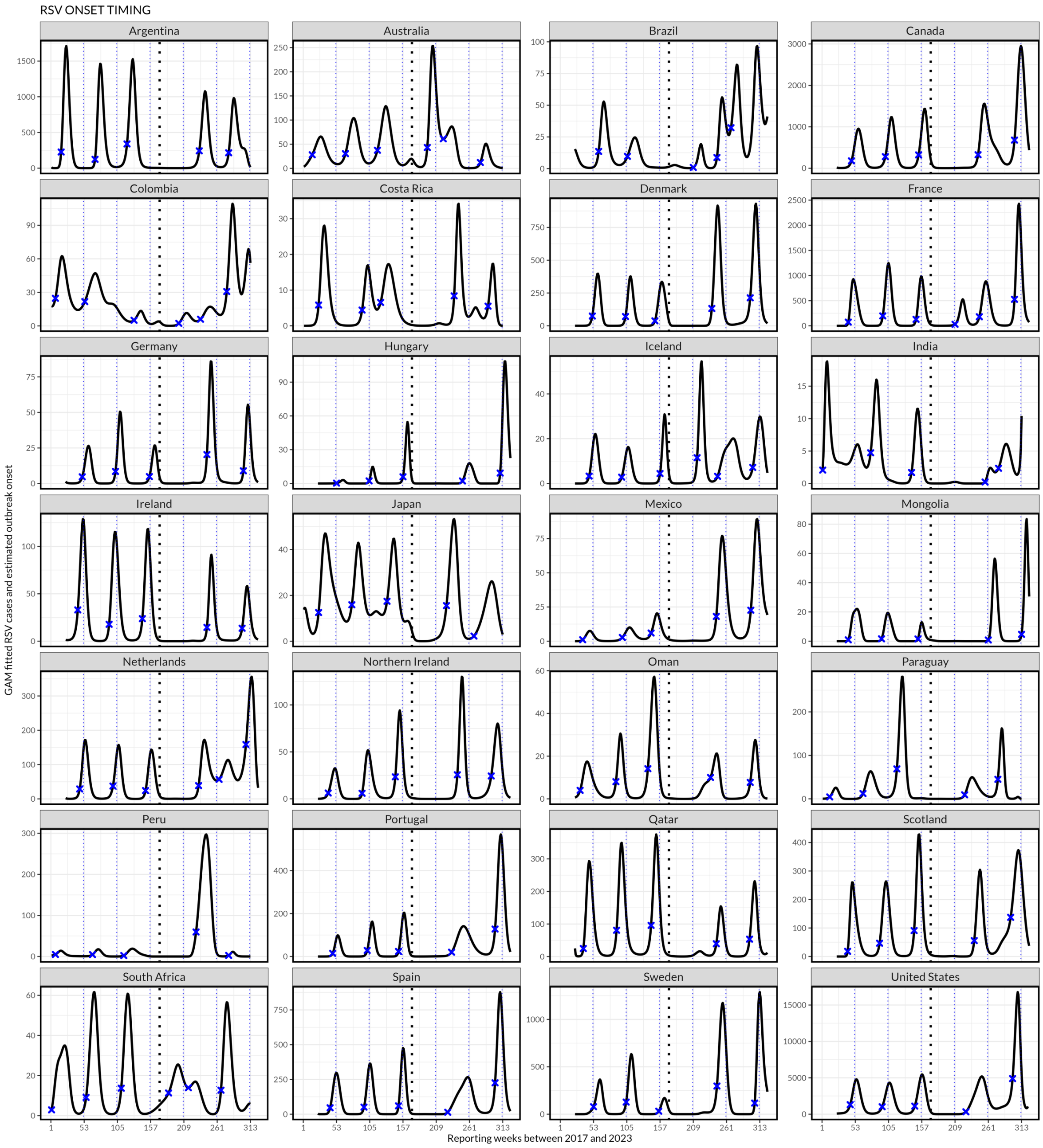


Supplementary Figure 1. Onset of respiratory syncytial virus (RSV) epidemics across 28 member countries of the World Health Organisation (WHO). The solid black line corresponds to the fitted P-spline, the dotted black line corresponds to April 2020 at the beginning of COVID-19 pandemic, and the blue star corresponds to the maximum second derivative value in the segment of increasing first derivative of the fitted P-spline GAM which defines the start of RSV epidemic (onset) as described in Figure 1.

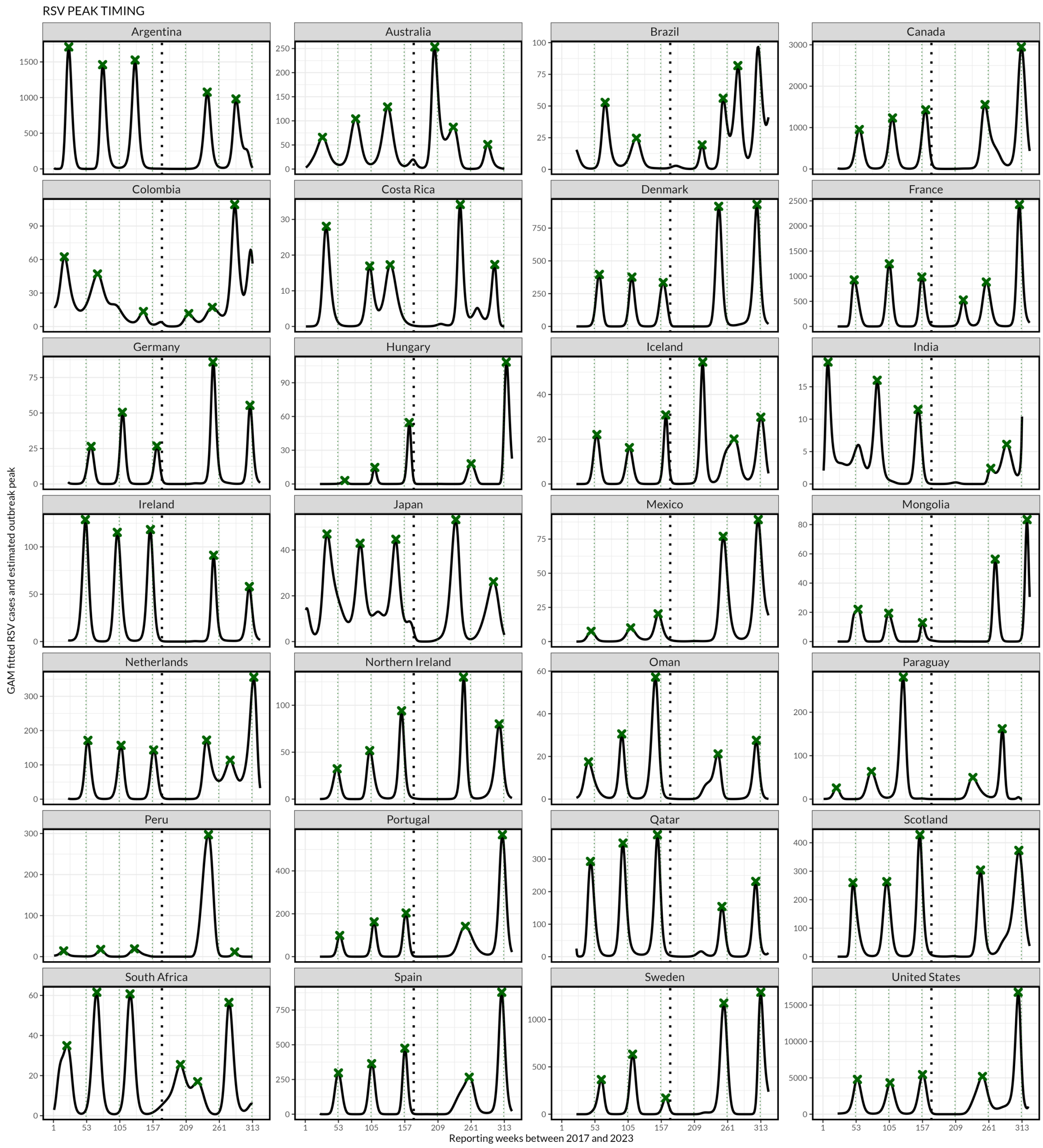


Supplementary Figure 2. Peak timing of respiratory syncytial virus (RSV) epidemics across 28 member countries of the World Health Organisation (WHO). The solid black line indicates the generalised additive model (GAM) fit with P-splines, the dotted black line corresponds to April 2020 at the beginning of COVID-19 pandemic, and the green star corresponds to the maximum value of the black fitted P-spline GAM, defining the wave peak of RSV cases.

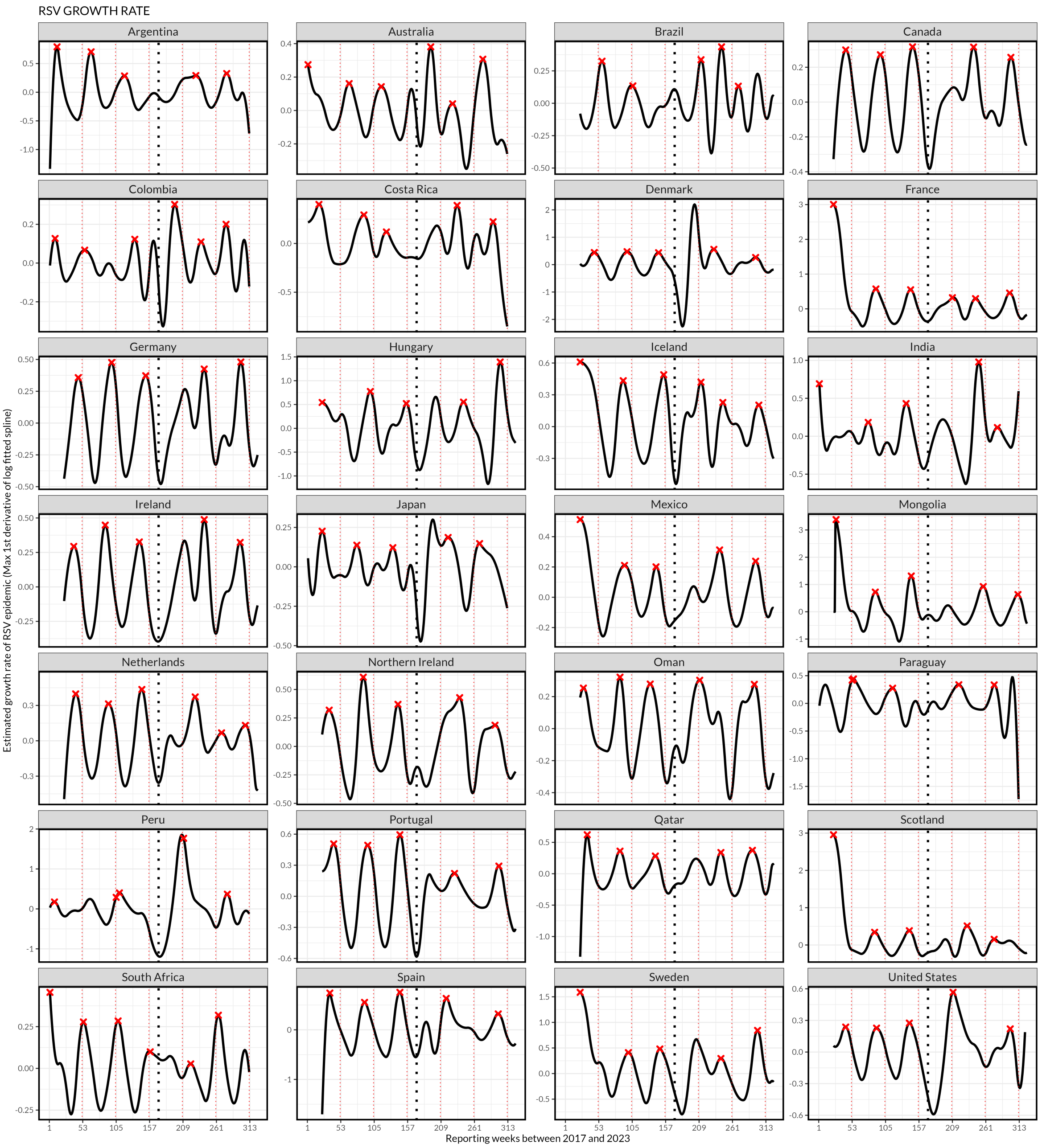


Supplementary Figure 3. Growth rate of respiratory syncytial virus (RSV) epidemics across 28 member countries of the World Health Organisation (WHO). The solid black line corresponds to the first derivative of the log of the fitted P-spline GAM, the dotted black line corresponds to April 2020 at the beginning of COVID-19 pandemic, and the red star corresponds to the maximum value of the black line which defines the maximum number of new cases per week (growth rate) as described in Figure 1.

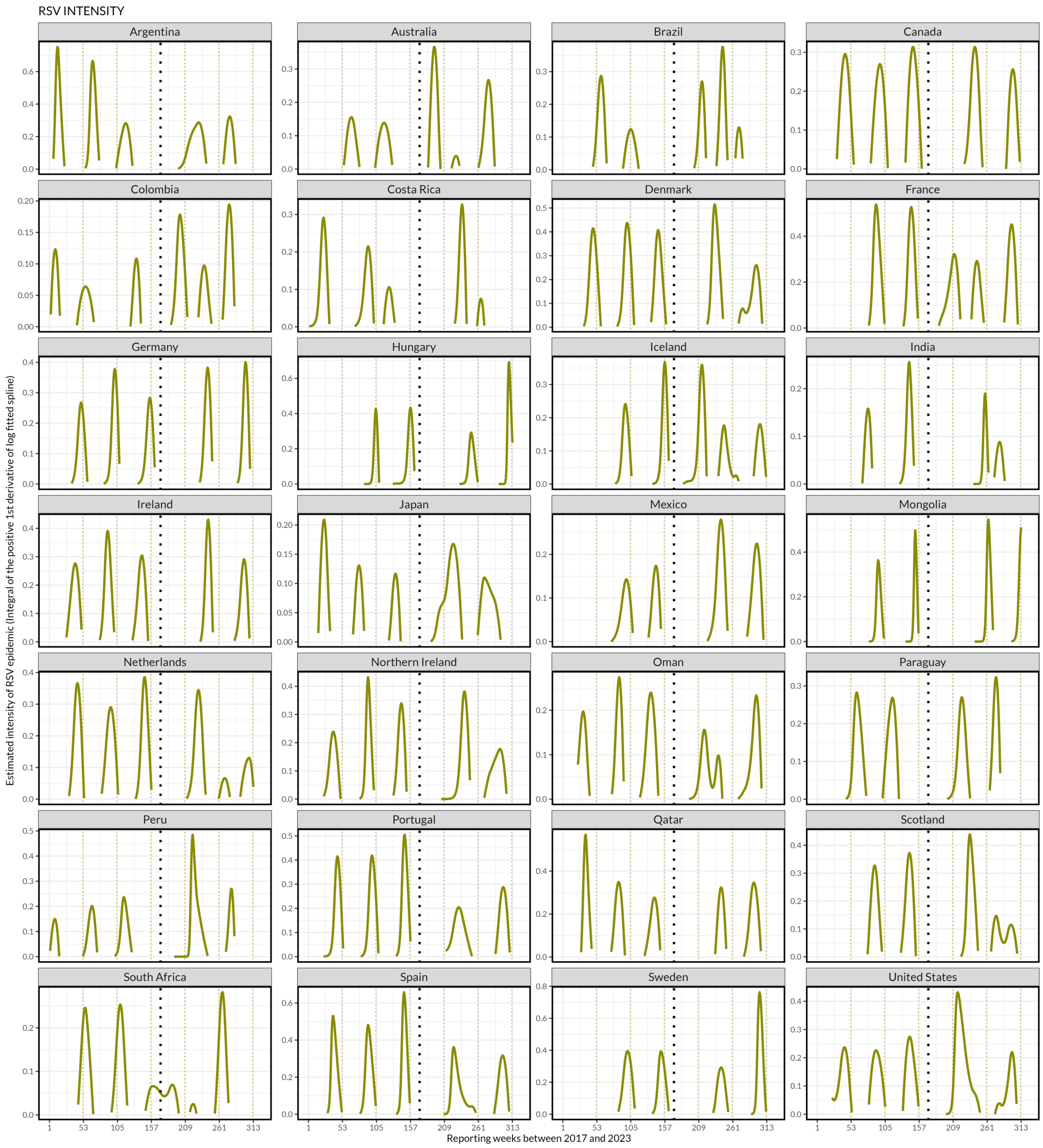


Supplementary Figure 4. Intensity of respiratory syncytial virus (RSV) epidemics across 28 member countries of the World Health Organisation (WHO). The dotted black line corresponds to April 2020 at the beginning of COVID-19 pandemic, and the area under the curve indicates the relative magnitude of RSV cases in each wave corresponding to the integral of the positive derivative of the log fitted black GAM P-spline before epidemic peak timing.

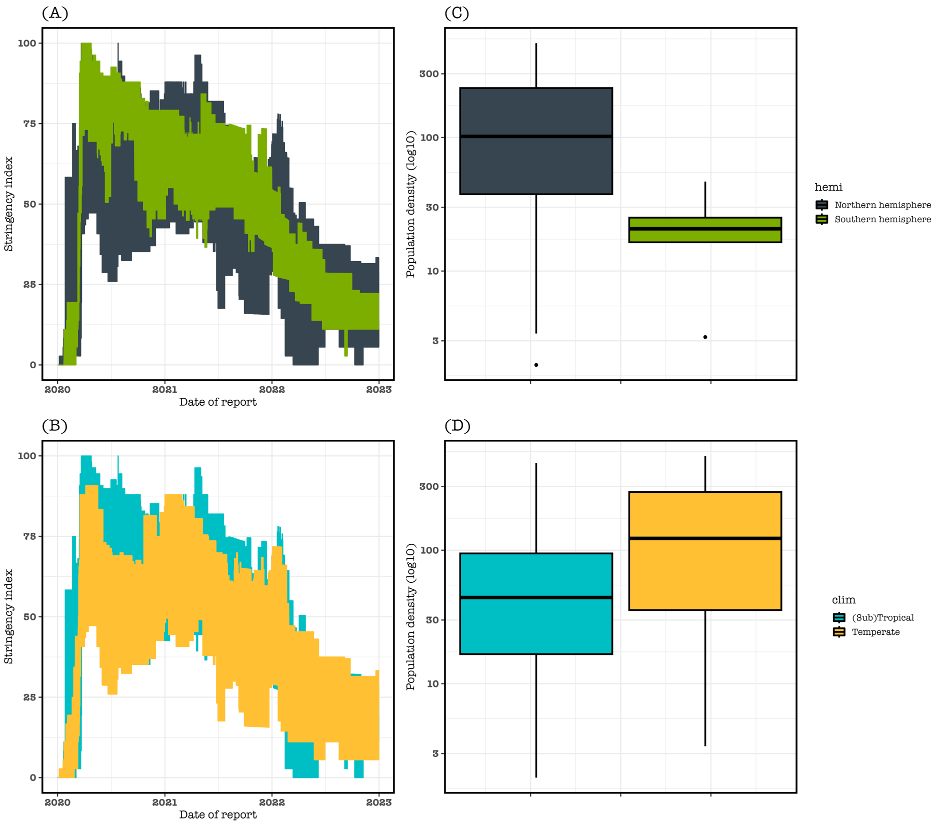


Supplementary Figure 5. Distribution of contact stringency index and population density by hemisphere and climate zone. (A, B) The contact stringency index is sourced from the Oxford COVID-19 Government Response Tracker data and uses nine metrics to calculate the Government Stringency Index including school closures, workplace closures, cancellation of public events, restrictions on public gatherings, closures of public transport, stay-at-home requirements, public information campaigns, restrictions on internal movements, and international travel controls. The stringency index is stratified by hemisphere and climate zone, with value of 0 referring to no restriction and 100 to maximum restrictions. (C, D) Box plot showing the spread of individuals per square kilometer (population density), in each hemisphere and climate zone.

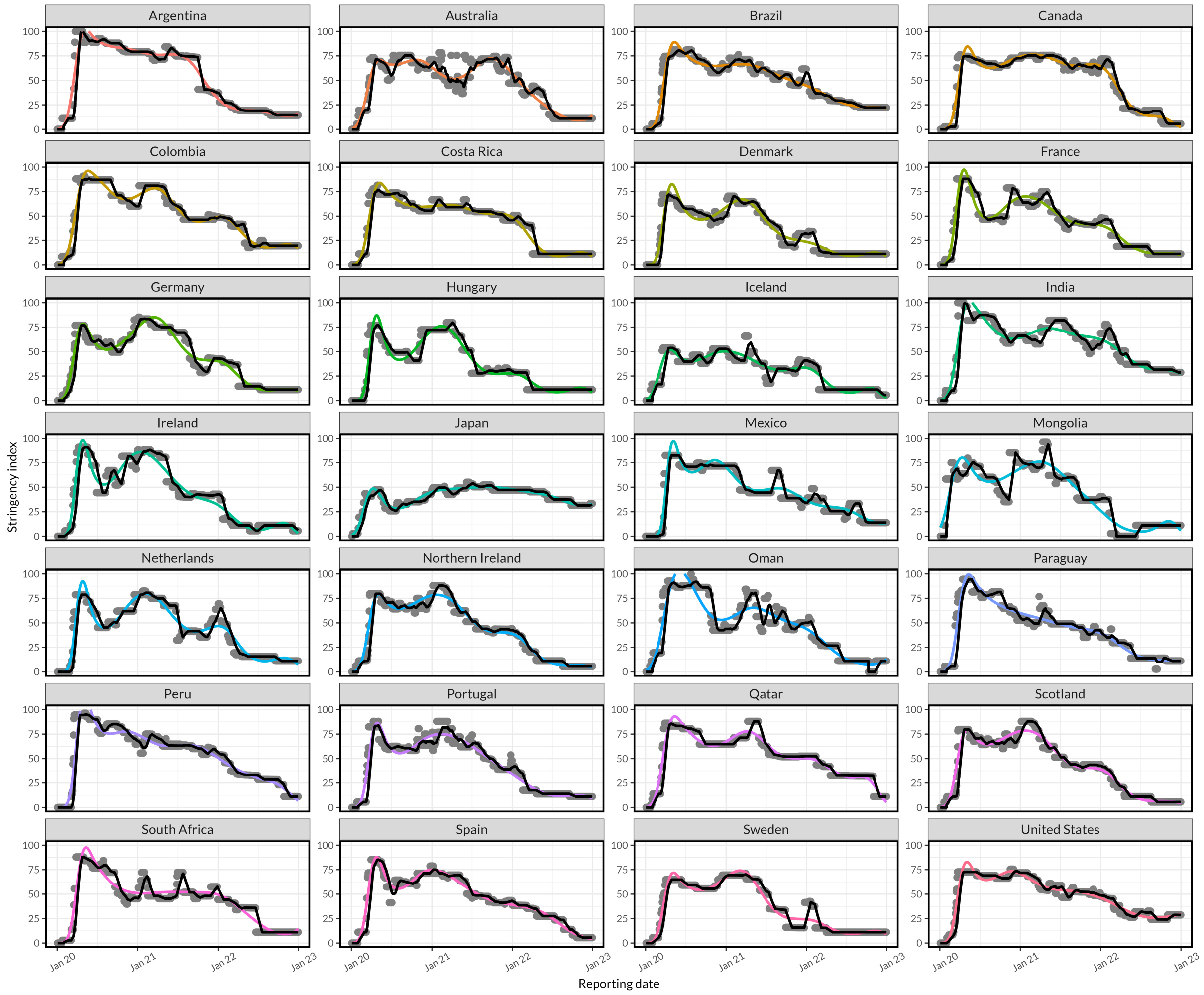


Supplementary Figure 6. Distribution of contact stringency index in 28 countries, globally. Thirty-days moving average (Black line), and generalised additive model (Color lines) fitted to reported stringency index (Gray points) to smooth out short-term effects of contact stringency. The contact stringency index is sourced from the Oxford COVID-19 Government Response Tracker data and uses nine metrics to calculate the Government Stringency Index including school closures, workplace closures, cancellation of public events, restrictions on public gatherings, closures of public transport, stay-at-home requirements, public information campaigns, restrictions on internal movements, and international travel controls. The stringency index has value of 0 referring to no restriction and 100 to maximum restrictions.

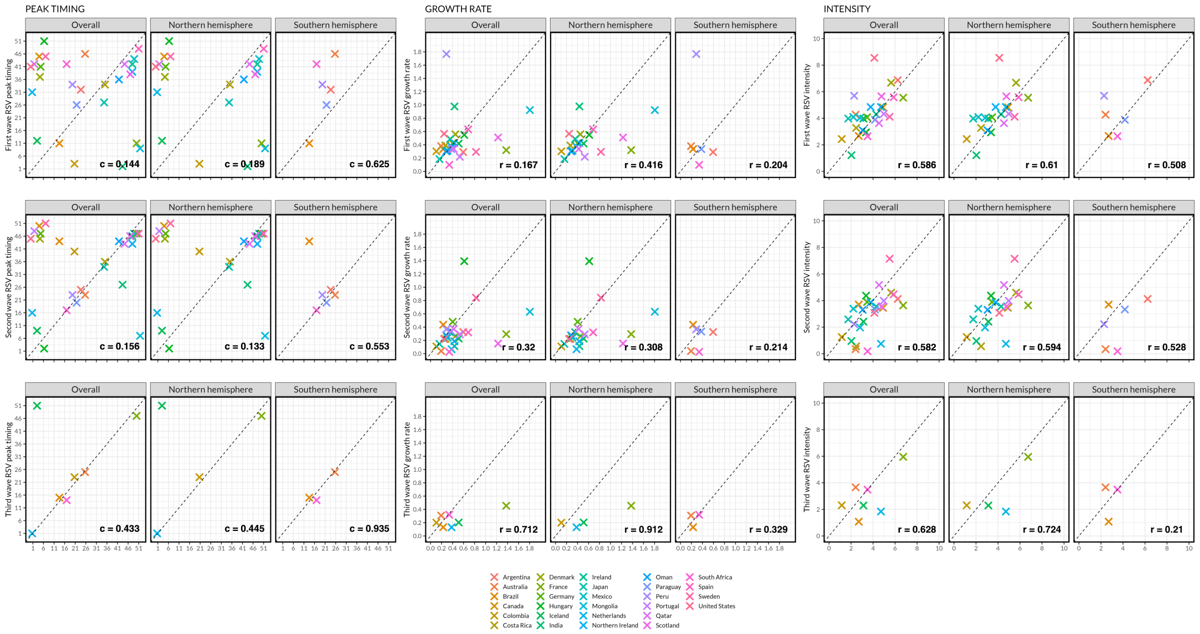


Supplementary Figure 7. Respiratory syncytial virus (RSV) epidemic peak timing, growth rate and intensity in 28 countries. Comparing the RSV epidemic peak timing, growth rate and intensity between pre COVID-19 vs first, second and third waves of RSV in all countries, and by northern and southern hemispheres. The $c$ metric refers to the circular correlation coefficient whereas $r$ metric is the Pearson's correlation coefficient.

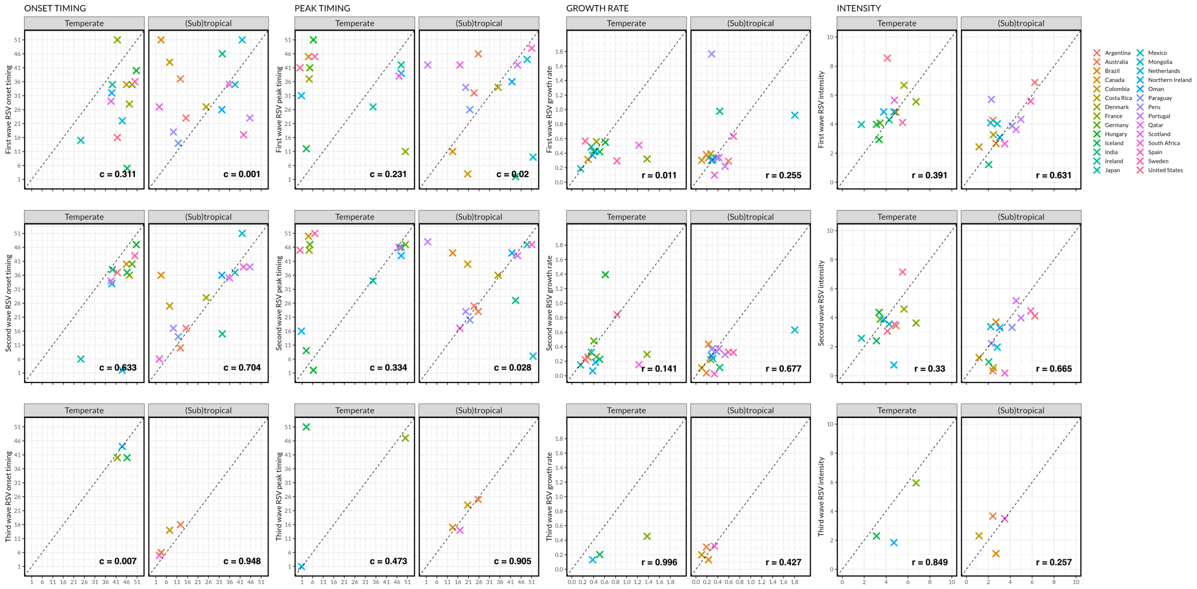


Supplementary Figure 8. Respiratory syncytial virus (RSV) epidemic onset timing, peak timing, growth rate and intensity in 28 countries. Comparing the RSV epidemic onset timing, peak timing, growth rate and intensity between pre COVID-19 vs first, second and third waves of RSV in all countries, and by Temperate and (Sub)tropical. The *c* metric refers to the circular correlation coefficient whereas *r* metric is the Pearson's correlation coefficient.

Supplementary Table 1. Correlation coefficients between different RSV waves post-COVID-19 and pre-COVID-19 mean values among countries that had a third wave of RSV.

| Season epidemic metric | Correlation between pre-COVID-19 and first wave of RSV | Correlation between pre-COVID-19 and second wave of RSV | Correlation between pre-COVID-19 and third wave of RSV |
|---|---|---|---|
| Onset† | 0.47 | 0.18 | 0.99 |
| Peak† | 0.11 | 0.68 | 0.43 |
| Growth rate‡ | 0.05 | 0.35 | 0.71 |
| Intensity‡ | 0.80 | 0.36 | 0.63 |

† Estimated with circular correlation coefficient

‡ Estimated with Pearson's correlation coefficient

Countries that had 3 waves of RSV following COVID-19 pandemic included Australia, Brazil, Colombia, France, Iceland, Netherlands, South Africa.

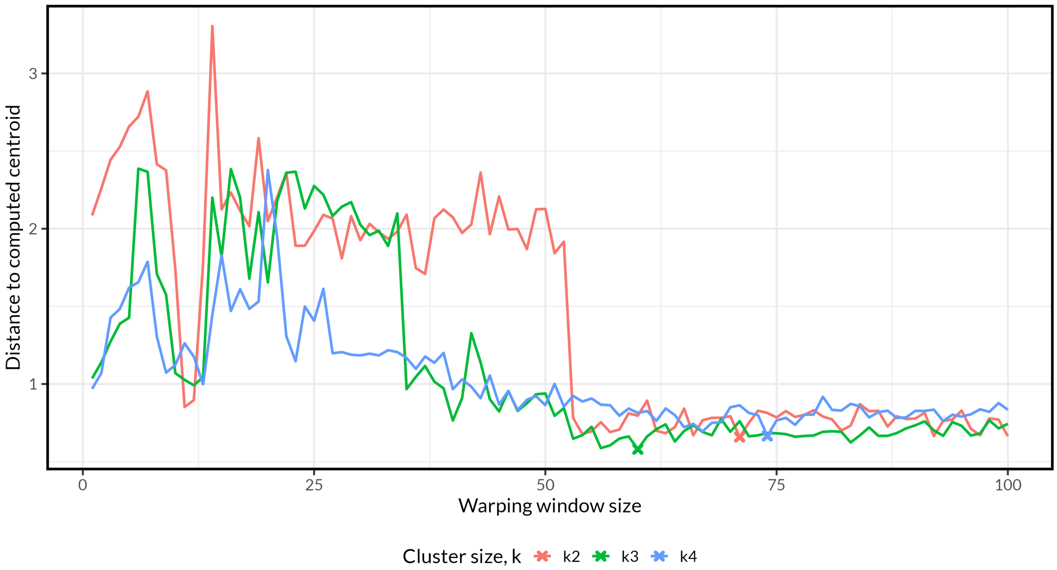


Supplementary Figure 9. Evaluation of the optimal DTW window size required to compute hierarchical clustering using Modified-Davies-Bouldin cluster validation index which is based on calculating the distances from time series to their centroid, where the optimal DTW window size is that which minimizes the distances. The centroid of each cluster is computed using DTW barycentre averaging function. The optimal DTW window size is identified at *w=71* for two clusters, *w=60* for three clusters, and *w=74* for four clusters.

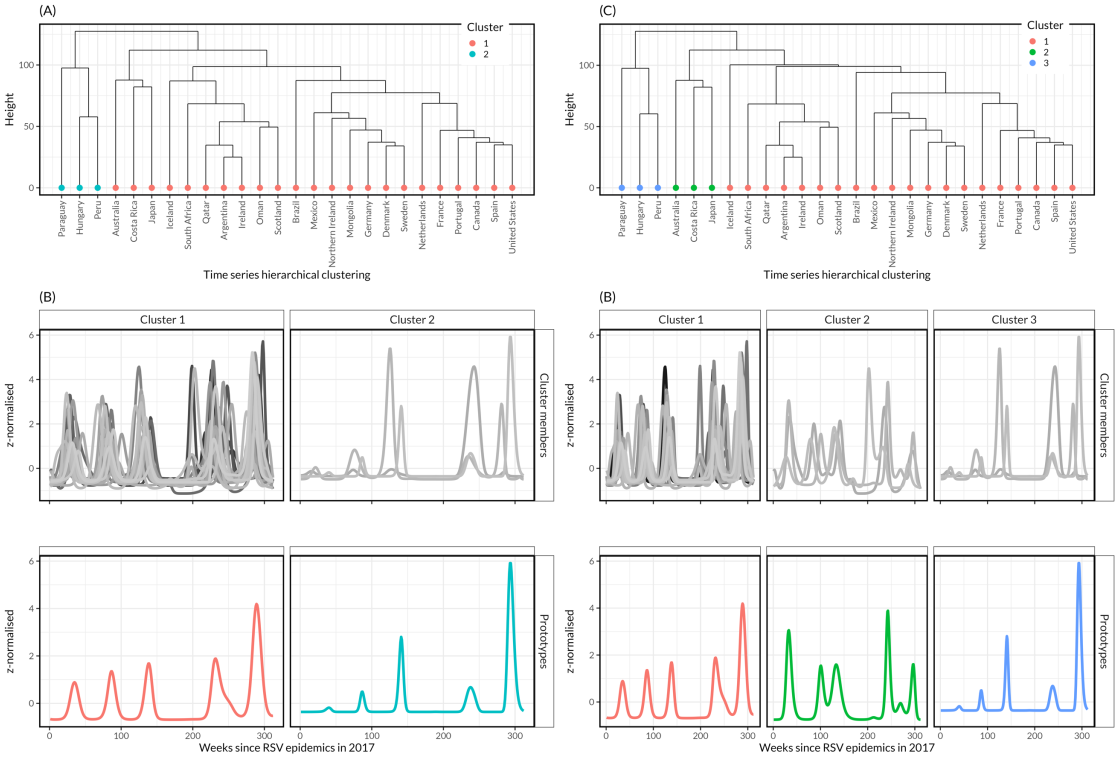


Supplementary Figure 10. Sensitivity plots of time series dynamic time warping and time series classification using hierarchical clustering. (A, B) Dendrogram representing hierarchical clustering and its corresponding series prototypes if the clusters size is set to 2 and warping window size to 71 as optimal combination of parameters based on Modified Davies-Bouldin (DB) internal cluster validity index (CVI). (C, D) Dendrogram representing hierarchical clustering and its corresponding series prototypes if number of clusters is set to 3 and warping window size to 60 based on DB and CVI.
